# Impact of COVID-19 on tuberculosis notifications in Blantyre Malawi: an interrupted time series analysis and qualitative study with healthcare workers

**DOI:** 10.1101/2021.03.15.21253601

**Authors:** Rebecca Nzawa Soko, Rachael M Burke, Helena RA Feasey, Wakumanya Sibande, Marriott Nliwasa, Marc YR Henrion, McEwen Khundi, Peter J Dodd, Chu Chang Ku, Gift Kawalazira, Augustine T Choko, Titus H Divala, Elizabeth L Corbett, Peter MacPherson

## Abstract

COVID-19 may impact on tuberculosis (TB) diagnosis and care. We analysed a city-wide electronic TB register in Blantyre, Malawi and interviewed TB officers. Malawi had no official “lockdown” but closed schools and borders on 23^-^March 2020. In interrupted time series analysis, there was an immediate 35.9% reduction in TB notifications (95% CI 22.0 to 47.3%) in April, which recovered to near pre-pandemic numbers by December 2020, but with 333 (95% CI 291 to 375) fewer cumulative notifications than anticipated. Women and girls were impacted (30.7% fewer cases, 95% CI 28.4 to 33.0%) more than men and boys (20.9% fewer, 95% CI 18.5 to 23.3). Fear of COVID-19 infection, temporary facility closure, inadequate protective equipment and COVID-19 stigma with similar presenting symptoms to TB were mentioned. Public health measures could benefit both TB and COVID-19, but only if diagnostic services remain accessible and are considered safe to attend.

## Introduction

Tuberculosis (TB) is a major killer, with approximately 1.4 million deaths annually (1) making it second only to COVID-19 as the biggest cause of infectious deaths in 2020.(2)

As well as the direct health effects of COVID-19, the secondary effects of the COVID-19 pandemic— including lockdowns, economic turmoil, healthcare worker illness and attrition, overwhelmed health facilities and fear of healthcare facilities—may impact on delivery of health services.(3) Concerns have been raised that COVID-19 may adversely impact on TB disease diagnosis, treatment and prevention, reversing recent progress in improving TB case detection and mortality, although physical distancing measures for COVID-19 could also reduce TB transmission.(1,3) Initial modelling, published in May 2020, suggested that healthcare service disruption worldwide could lead to 6.3 million additional cases of TB and an additional 1.4 million TB deaths between 2020 and 2025 due to under-diagnosis of TB and TB treatment interruptions.(4) Empirical data from high TB burden settings is urgently needed to better understand the impact of COVID-19 on TB, and to inform mitigation strategies.

Malawi is one of thirty high TB/HIV burden countries.(1) In Blantyre, located in the Southern Region Malawi, a citywide electronic TB register has been maintained in partnership by Malawi Liverpool Wellcome Trust, the Malawian National TB Programme, and Blantyre District Health Office.(5) We used these data to investigate the impact of COVID-19 on citywide TB case notifications. We hypothesised that the direct and indirect effects of COVID-19 epidemic in Malawi would reduce TB case notifications and this may have been disproportionately experienced at different health system levels and by certain population groups, including people living with HIV. Our primary objective was to estimate the number of missed TB case notifications, and our secondary objective was to determine whether this was affected by sex, health facility or HIV status. Finally, in order to investigate and explain the underlying causes of under-notification of TB, we undertook a qualitative study with TB Officers, the cadre of healthcare worker who provide most TB services in Malawi.

## Methods

### Data sources

To estimate population denominators for Blantyre District, we obtained age- and sex-specific background mortality rates and fertility rates from 2008-2020 World Population Prospect (WPP) data (6) and used the Cohort-Component method to combine them into local estimates from the 2008 and 2018 Malawi National population censuses.

In Blantyre, TB Officers working at all primary health centres and the city’s main central hospital (Queen Elizabeth Central Hospital [QECH]) record demographic and clinical characteristics of all TB patients who register for treatment using an electronic case record form. Data collected includes date and clinic of registration, age, sex and HIV status, residential address, and TB characteristics (pulmonary vs. extra-pulmonary and microbiological classification). Records are reconciled with Ministry of Health National TB Programme treatment registers every quarter. Each month, a randomly-selected 5% sample of registered TB cases undergo home tracing for data validation purposes.

### Statistical modelling

To investigate the impact of COVID-19 on TB case notification in Blantyre we conducted an interrupted time series analysis. (7) The Malawi Government declared a state of emergency due to COVID-19 on the 23^rd^ March 2020, and the first COVID-19 cases were diagnosed on 2^nd^ April 2020. We assumed that implementation of COVID-19 restrictions and the government and public response to the emerging epidemic would cause both an immediate “step change” change in TB case notifications and a “slope change” leading to different month-by-month trends than seen pre-COVID.(7) Using a negative binomial distribution to account for overdispersion, we modelled monthly counts of TB cases as a function of month, COVID-19, and month-given-COVID (Model 1). We used notification data from June 2016, as this marks the start of Malawi introducing “test and treat” for antiretroviral therapy (ART) provision and the implementation of Xpert for TB diagnosis.

## Model 1

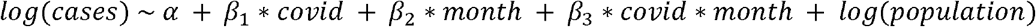

Where cases=expected numbers of notified TB cases per month, ⍰=intercept, covid=indicator variable for period before or after introduction of COVID restrictions (before or after 1^st^ April 2020), month=indicator for month from June 2016, and population=monthly updated population denominator for Blantyre District

We estimated trends in TB case notification rates (CNRs) by converting model-fitted monthly numbers of notified cases to annualised equivalent cases per 100,000 population using estimated Blantyre census population denominators. We used the model to predict TB CNRs from April 2020 onwards under the counterfactual situation where COVID had not occurred and background trends (i.e. trend from April 2016 and March 2020) had continued linearly. We defined numbers of “missed” TB cases as the difference between the observed numbers of notified cases and numbers expected under the counterfactual no-COVID-19 situation (acknowledging that some of the “missed” cases may be diagnosed later and so be delayed rather than entirely missed). Ninety-five percent confidence intervals for the total number of “missed” TB cases were estimated through 1,000 parametric bootstrap replications. Observed cases are taken as is and predicted cases under the counterfactual are from a normal distribution (on the link scale) with mean equal to model prediction for given month under the counterfactual and standard deviation equal to model standard error for predictions for the given month under the counterfactual.

For the secondary objective, we modelled the differential impact of COVID on TB case notifications by sex, HIV status and whether TB was diagnosed at the Central Hospital or primary care level (Model 2). Because there was a small amount of missing data for HIV status and sex, we preformed multiple imputation using chained equations with predictive mean matching, using the R ‘mice’ package.(8)

### Sensitivity analysis

TB exhibits seasonality related to climate and weather conditions.(9) We therefore performed a sensitivity analysis by adding seasonal effects to the interrupted time series model using a harmonic term with two peaks every 12 months.

### Qualitative analysis

Between 21 October and 14 December 2020, in-depth interviews were conducted with 12 TB Officers from the Blantyre Health Facilities (2 from QECH, and 10 from primary health care centers) to gain an understanding of the main reasons for the changes in TB case notifications in the COVID-19 era. A local social scientist with experience of qualitative interviewing conducted interviews in Chichewa. Data were recorded and simultaneously transcribed and translated to English. A thematic framework was developed from the initial four interviews and applied across all subsequent interviews. Coding and data analysis were done using NVIVO. Interviews were continued until saturation of themes was reached.

### Ethical approval

Participants provided oral consent for their data to be recorded in the enhanced surveillance dataset; a waiver of requirement for written consent was approved by London School of Hygiene and Tropical Medicine and College of Medicine, University of Malawi, both of whom provided ethical approval for the Blantyre enhanced TB surveillance system and qualitative interviews. Participants in the in-depth interviews provided informed written consent.

### Role of funding source and data availability

The Blantyre Enhanced TB Surveillance data is funded by Wellcome, through a grant to ELC (200901/Z/16/Z). Wellcome had no role in design or conduct or this work, or the decision to publish. Data and code to recreate analyses are freely available at https://github.com/rachaelmburke/tbcovidblantyre.

## Results

### Interrupted time series

Between June 2016 and December 2020, 10,274 TB cases were notified in Blantyre. Between June 2016 and March 2020 (i.e. before COVID), annualised Blantyre TB CNRs fell by approximately 1% per month, with a peak of 405 cases per 100,000 people in November 2016, declining to 137 cases per 100,000 people in October 2019. A total of 9,199 people with TB were notified in Blantyre during this period: 3,561 among women and girls and 5,611 in men and boys (27 missing sex). People living with HIV represented 5,820 (63.3%) TB notifications and HIV-negative people made up 3,279 (35.6%) of the notified people with TB (100 people with missing or unknown HIV status). TB notifications were split fairly evenly between QECH (4,889 notifications, 53.1%) and primary health facilities (4,310 notifications, 46.9%). Children (aged ≤ 14 years old) comprised 920 (10%) of notifications. The median (interquartile range) age among adults diagnosed with TB was 35 (28 - 44) years for women and 37 (30 - 45) years for men.

The declaration of a national COVID-19 disaster led to an abrupt decline in TB notifications in April 2020 by 35.9% (95% CI 22.1 to 47.3%) – Figure 1. However, subsequently, TB notifications increased at a rate of 4.40% per month (0.59 to 8.36%). The effect of this initial decline at the start of the COVID pandemic was that observed Blantyre TB annualised CNRs in March 2020 (pre-COVID-19) and April 2020 (post COVID-19) were 240 and 152 cases per 100,000 people, respectively. This compares to a predicted April CNR of 230 cases per 100,000 person years in the counterfactual scenario with no COVID. However, by November and December 2020 observed Blantyre TB CNRs were 205 and 156 cases per 100,000 person-years, compared to a predicted CNR of 213 and 211 cases per 100,000 in the counterfactual scenario with no COVID.

**Figure 1:**
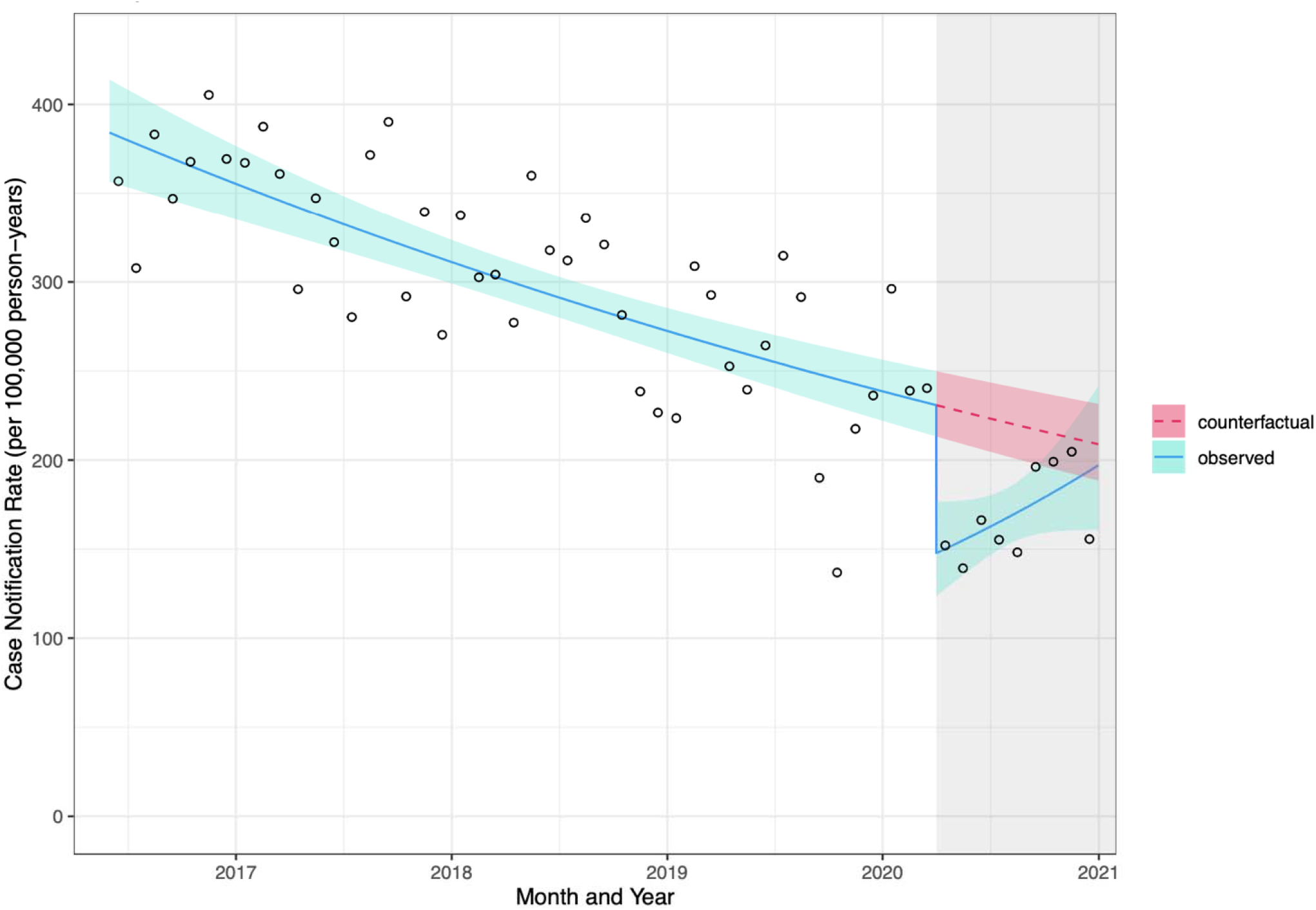
**Impact of COVID-19 on TB monthly tuberculosis case notification rate in Blantyre, Malawi** Dots = observed number of cases. Line = fitted model (95% CI) with both step and slope change due to COVID, see methods for details. Shaded areas indicate time that COVID emergency was declared in Malawi

Between April 2020 and December 2020, 1,075 cases of TB were notified in Blantyre (equivalent to 196 cases per 100,000 person-years) – Table 1. Under the counterfactual situation of no COVID-19 epidemic, we would have expected that 1,408 people with TB would have been notified (95% CI 1,366 to 1,451 cases, equivalent to annualised case notification rate of 221 cases per 100,000 people). We therefore estimate that the COVID epidemic (directly and indirectly) led to 333 fewer TB notifications (95% CI 291 to 376), equivalent to a 23.7% reduction in TB notifications (95% CI 21.4 to 26.0%).

**Table 1:** **Modelled impact of COVID-19 on Blantyre TB case notifications between April 2020 and December 2020**

|  | **Observed**<br>**numbers of**<br>**notified TB**<br>**cases (with**<br>**COVID-19)** | **Model-estimated**<br>**number of**<br>**notified TB cases**<br>**(counterfactual,**<br>**without COVID-**<br>**19)** | **Absolute**<br>**difference** | **Relative difference (%)** |
| --- | --- | --- | --- | --- |
| **Model 1** |  |  |  |  |
| Overall | 1075 | 1408<br>(1366 to 1451) | 333<br>(291 to 376) | 23.7<br>(21.4 to 26.0) |
| **Model 2** |  |  |  |  |
| Male | 692 | 875 (848 to 901) | 183 (156 to 209) | 20.9 (18.5 to 23.3) |
| Female | 383 | 553 (534 to 571) | 170 (151 to 188) | 30.7 (28.4 to 33.0) |
| Primary health centres | 488 | 761 (737 to 785) | 273 (249 to 297) | 35.9 (33.9 to 37.9) |
| Queen Elizabeth<br>Central Hospital | 587 | 666 (645 to 688) | 79 (58 to 101) | 11.9 (9.10 to 14.7) |
| HIV-positive | 660 | 820 (796 to 845) | 160 (136 to 185) | 19.6 (17.2 to 21.9) |
| HIV-negative | 415 | 607 (586 to 627) | 192 (171 to 212) | 31.6 (29.3 to 33.8) |

As a secondary objective, we modelled which population groups were most affected by disruption to TB services (Figure 2). This model incorporates sex, HIV status and healthcare facility (QECH vs. primary care clinics), and estimates slightly higher numbers of missed TB notifications (352, 95% CI 319 to 385) between April to December 2020. Men and boys accounted for a slightly larger number of missed TB diagnoses with 183 fewer notification (95% CI 158 to 209) compared to 170 fewer notifcations among women and girls (95% CI 151 to 188). However, women and girls had a larger proportional decline than men and boys (30.7%, 95% CI: 28.4 to 33.0%; compared to 20.9%, 95% CI 18.5 to 23.3%). Notifications at Primary healthcare clinics were also disproportionately reduced compared to hospital notifications, as were notifications for HIV-negative patients compared to those among people living with HIV (Table 2). Sensitivity analysis around seasonality of TB did not materially affect the conclusions

**Table 2.**
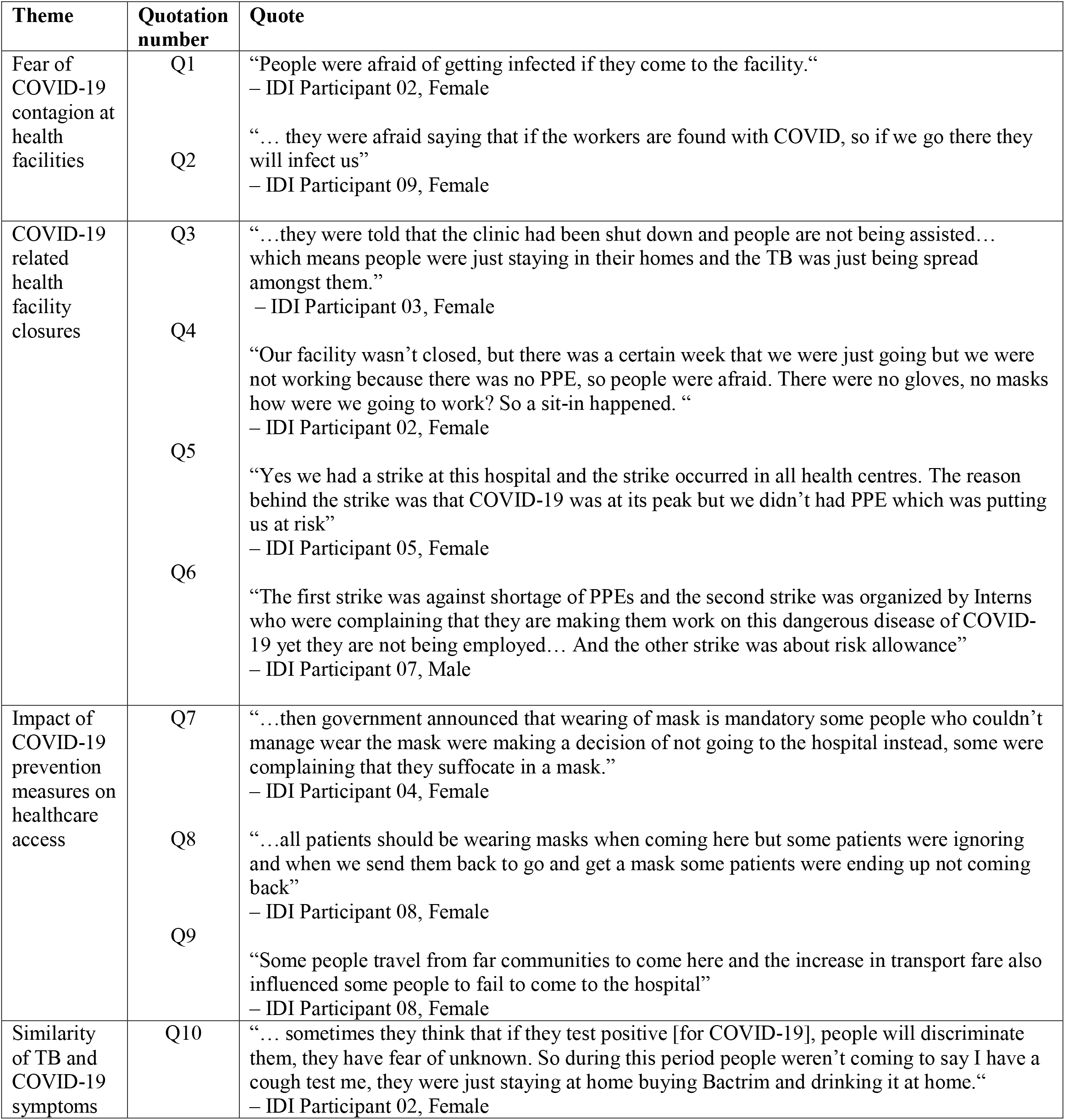
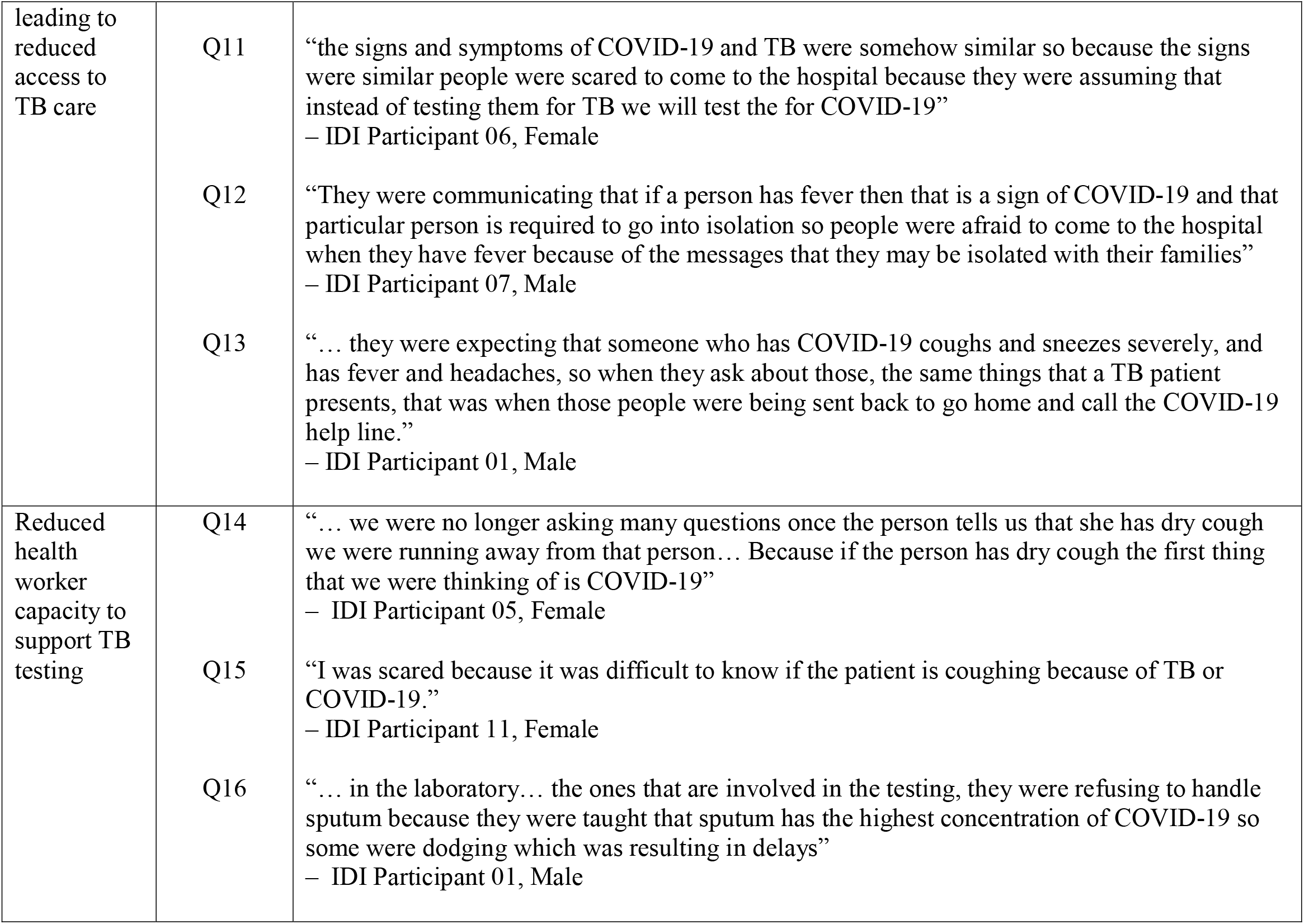
**Quotations from TB Officers in-depth interviews about reasons for reduced TB notifications.**

**Figure 2:**
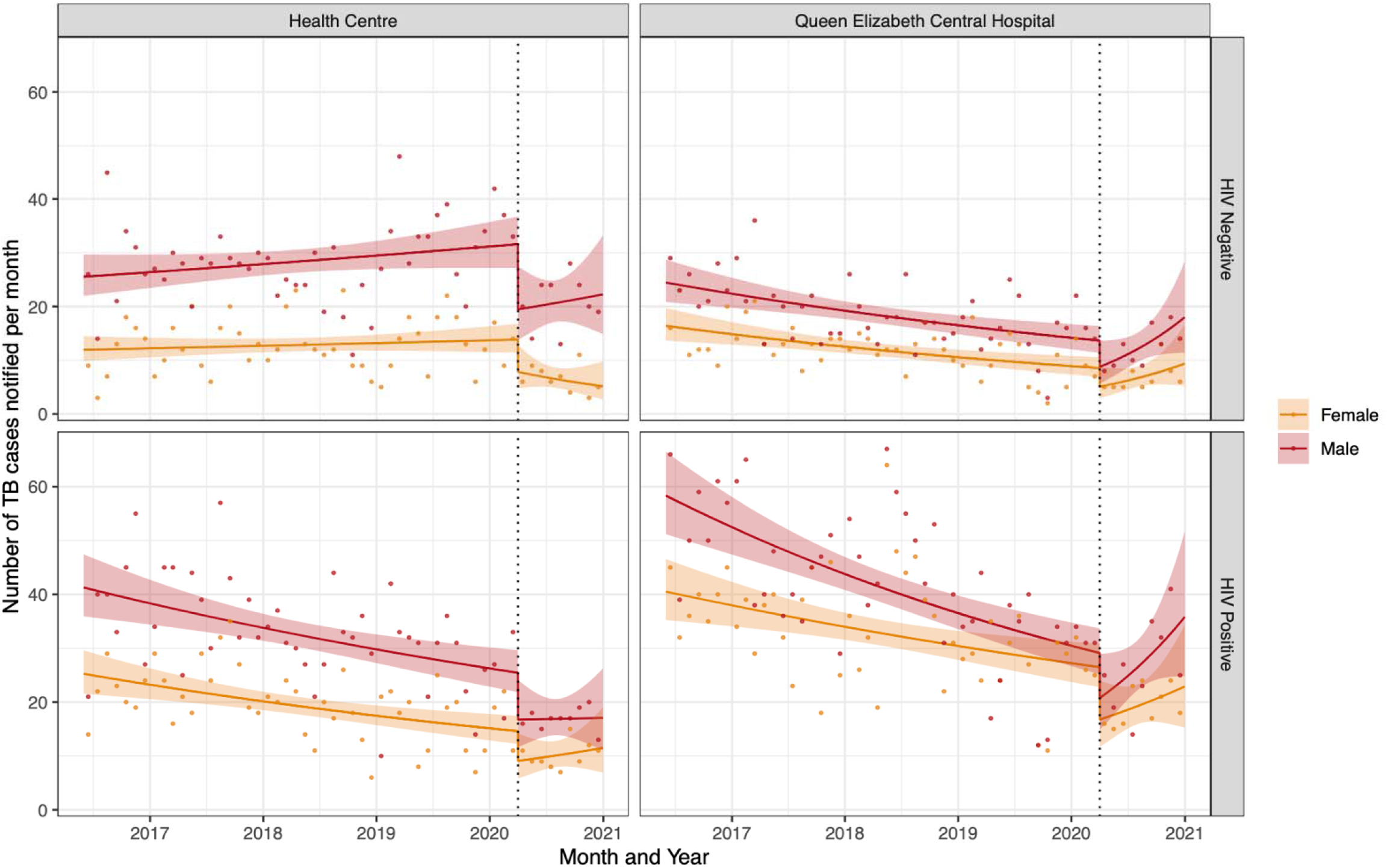
**Impact of COVID-19 on TB case notifications in Blantyre, Malawi by HIV status, registration site and sex** Dots = observed number of cases per month. Line = fitted model (95% CI) with both step and slope change due to COVID, see methods for details. Vertical lines indicate time that COVID emergency was declared in Malawi

### Qualitative results

Of the 12 in-depth interviews with health providers, nine participants were female, and three were male, with ages ranginh from 34 to 53 years. Ten had secondary-level education.

Themes that emerged from the in-depth interviews related to both an overall reduction in people attending health facilities and to a reduction in quality of TB services provided. Reduced attendance affecting all services at health facilities were said to reflect a combination of fear among general public of getting infected with COVID-19 if they attended the facility (fear of contagion), closures related to disinfection following health-worker diagnosis with COVID-19 and health-worker strikes, and COVID-19 prevention measures such as face masks and social distancing that lead to reduced capacity at any given clinic and hence access to care. TB specific issues included the overlap between common TB symptoms and symptoms of COVID-19. Fearfulness among general clinic and TB service staff together with initial shortages in PPE left providers unable to support sputum submission for TB testing, so that people with those symptoms were instead turned away from facilities lacking personal protective equipment (PPE).

### Reduced attendance at health facilities

As well as reduced attendance from fear of becoming infected with COVID-19 among the general public, several health workers tested positive for COVID-19 during the epidemic (quote 1 & quote 2, Table 2). These facility based COVID-19 outbreaks led to temporary facility closure for disinfection. This not only affected numbers attending the health facilities on the days of closure but also led to greater fear of infection there and in one instance rumours that the clinic was closed for a longer period of time (quote 3, Table 2). Finrally, strikes and ‘sit-ins’ over both risk allowance payments and lack of (PPE) also resulted in temporary closures of facilities (quote 4 – quote 6, Table 2)

### Regulations about facemasks and public transport

Government COVID-19 prevention measures on face masks and social distancing requirements were also reported to have contributed to reduced access to health services. Mandatory use of face masks at health facilities was introduced during the epidemic, but TB officers cited the inability to afford a mask and the feeling that masks “suffocate them” as reasons for patients not wanting to wearing one (quote 7, Table 2). Patients who tried to attend facilities without having a mask were “sent back” (meaning that they were not seen by a healthworker) and often did not return (quote 8, Table 2). Public transportation in Blantyre also had a limit on vehicle capacity, which led to doubled transport costs, limiting clinic access (quote 9, Table 2).

### TB specific issues

Since the presenting symptoms for TB and COVID-19 are very similar (both cough and fever), TB officers reported this led to particular issues around TB testing. Firstly, people with these symptoms were reportedly afraid of being tested for COVID-19 if they attended the facilities. TB Officers said they were more afraid of COVID-19 than TB because they knew that TB could be cured (quote 10 & quote 11, Table 2) and also that patients with COVID-19 may need to be placed under facility isolation (quote 12, Table 2). This similarity of symptoms also led to people who would normally have been tested for TB being turned away from the facility and told to instead “go home and call the COVID help line” (quote 13, Table 2).

The TB officers themselves also spoke of their own fear of contracting COVID-19 infection from presumptive TB patients, given the similarity of symptoms. They changed how they interacted with symptomatic people, including interacting less directly, and not supervising sputum collection as closely (quote 14 & quote 15, Table 2). In addition, a lack of personal protective equipment in health facilities was also reported to have forced most TB officers to temporarily stop conducting TB tests or supervising sputum collection at all. For those patients who did submit sputum results could be delayed due to fears of laboratory staff that the sputum may contain high levels of COVID-19 (quote 16, Table 2).

## Discussion

As well as directly causing millions of deaths, the COVID-19 pandemic has directly and indirectly impacted on delivery of health services throughout the globe.(10) In this analysis of the impact of the COVID-19 pandemic on TB notifications in Blantyre Malawi, we found a substantial immediate decline in TB case notifications concurrent with the start of the COVID-19 pandemic in Malawi. This is consistent with initial reports on COVID impact from other settings.(11–18) However, we show that following the initial decline, TB case notification rates then increased and reached near pre-pandemic levels within nine months. Overall, we estimate that there were 333 fewer cases of TB notified (equivalent to 39 per 100,0000 people) from April 2020 to December 2020 than would have been expected if there was no COVID-19 epidemic. For the affected individuals these missed or delayed diagnoses are likely to have severe consequences, and for public health programmes the consequences might hinder progress towards eliminating TB. The TB case notifications may also be indicative of more general disruption of a range of primary healthcare services.

To put these results into context, Malawi is a high HIV/ TB burden country. Estimated prevalence of TB in urban Malawi was 988 per 100,000 people at the last national survey in 2013.(19) TB in Malawi is declining, due to concerted efforts from the national and District TB and the HIV programmes. Test and treat for HIV (i.e. starting antiretroviral therapy regardless of CD4 cell count) was introduced in Malawi in June 2016 and Malawi is coming close to achieving UNAIDS HIV 90-90-90 goals. However, TB remains one of the leading causes of death and years of life lost in Malawi.

We consider that the major reason for the drop in notifications is that people with true TB disease had their TB diagnosis missed, or at least delayed. This is consistent with data from our qualitative interventions with TB officers, who noted that—in the immediate period after the start of the Malawi COVID-19 epidemic—access to health facilities was extremely challenging. Alternative explanations are that people were diagnosed with TB and started on treatment but not notified to the national programme, or that the true incidence of TB declined. We consider these explanations unlikely. TB treatment cannot be accessed in Malawi outside of TB registration centres, and our electronic TB surveillance system is cross-referenced with paper ledgers that confirm the same trends in notifications. Reduced incidence of other respiratory pathogens, notably influenza, have resulted from the non-pharmaceutical interventions for COVID-19, and it is possible that TB transmission has also declined. However, the prolonged interval between infection and onset of symptoms for TB makes an immediate impact on notifications within three months implausible, particularly as Malawi has had less stringent COVID-19 prevention measures than many other countries.

Our qualitative interviews indicate that in addition to general restrictions on healthcare access during the COVID-19 epidemic TB testing and notifications were particularly affected due to the similarity in presenting symptoms between TB and COVID-19. Not only were people with TB symptoms less likely to attend facilities for fear of COVID-19 diagnosis and possible consequences such as isolation, but those who did attend were likely to be turned away and directed to COVID-19 specific services where they would be unlikely to be assessed for TB. In high TB burden countries alignment of COVID-19 and TB diagnosis, prevention and care is likely to lead to improved outcomes for both diseases.

Women had disproportionately higher reductions in case notifications than men, as did HIV-negative compared to HIV-positive patients, and notifications from primary care clinics compared to the central hospital. We hypothesise that women faced greater barriers to accessing healthcare during COVID than men, possibly due to greater requirements to home-school children, or social gender norms meaning that men were more likely to disregard COVID-19 public health restrictions, and perhaps additionally due to economic requirements to leave the house to work.(20)

Primary healthcare centres were more affected than the central hospital, both in terms of initial step change (drop in TB cases notified at the start of COVID-19) and with slower recovery in the period after COVID-19 Reasons for this could include the central hospital being prioritised for personal protective equipment and so remaining more fully functional. Patients diagnosed with TB at the hospital tend to have more severe illness, and so potentially were unable to delay healthcare seeking. TB cases among HIV negative people declined more than among people living with HIV. This could be associated with site of TB diagnosis: the central hospital has the by far largest number of HIV-positive people registered for ART in the city. Alternatively, PLHIV may have more severe TB symptoms and be less able to defer healthcare seeking.

Limitations to this work include uncertainty around the counterfactual conditions; between June 2016 and March 2020 TB case notifications were declining and for the counterfactual “no COVID” scenario we modelled them as continuing to decline at the same rate. Since December 2020, Malawi has had a second wave of COVID-19. Whilst our electronic enhanced surveillance data is entered in real time, data is monitored and verified on a quarterly basis, so we do not yet have information on the impact of the COVID-19 second wave in Malawi.

Malawi is also fortunate to have well-functioning TB and HIV programmes that may be more resilient to COVID-19 than in other countries. Malawi did not introduce any substantial restrictions on population movement and gathering due to COVID-19, so there have been no legal restrictions on travelling to TB clinics. Therefore, our data are not necessarily generalisable to other settings in Southern Africa or elsewhere.

The individual and household impact of missed or delayed TB diagnoses are likely to be severe for the affected people. However, it is relatively reassuring that the initial COVID-19 related decline in TB case notification was not sustained and that the Malawi TB programme had a relatively quick recovery once COVID-19 first wave rescinded. We observed a shorter period of disruption that earlier modelling of COVID-19 impact on TB assumed.(4) It is neither possible nor desirable to consider either COVID-19 or TB diagnosis, treatment, care and public health measures in isolation. Rather there may be opportunities to combine resources to tackle both COVID-19 and TB, through improved infection prevention and control at health facilities, strengthened laboratory infrastructure and community engagement to address stigma and provide sources of information about both diseases, in a setting of universal health coverage.

## Data Availability

Data and code to recreate analyses are freely available at https://github.com/rachaelmburke/tbcovidblantyre

## Acknowledgements

The Blantyre Enhanced TB Surveillance data is funded by Wellcome, through a grant to ELC (200901/Z/16/Z). RMB and PM are funded by Wellcome (203905/Z/16/Z and 206575/Z/17/Z, respectively). MYRH is supported by a Wellcome Strategic Award to the Malawi Liverpool Wellcome Trust Clinical Research Programme (206545/Z/17/Z). PJD was supported by a fellowship from the UK Medical Research Council (MR/P022081/1); this UK-funded award is part of the European and Developing Countries Clinical Trials Partnership 2 (EDCTP2) programme supported by the EU. All authors declare no conflicts of interest.

## Notes

### Competing Interest Statement

The authors have declared no competing interest.

### Author Declarations

London School of Hygiene and Tropical Medicine Research Ethics Committee University of Malawi College of Medicine Research Ethics Committee.

